# Mechanical ventilation utilization in COVID-19: A systematic review and meta-analysis

**DOI:** 10.1101/2020.06.04.20122069

**Authors:** Mohammed A. Almeshari, Nowaf Y. Alobaidi, Mansour Al Asmri, Eyas Alhuthail, Ziyad Alshehri, Farhan Alenezi, Elizabeth Sapey, Dhruv Parekh

## Abstract

**Background:** In December 2019, SARS-CoV-2 caused a global pandemic with a viral infection called COVID-19. The disease usually causes respiratory symptoms but in a small proportion of patients can lead to a pneumonitis, Adult Respiratory Distress Syndrome and death. Invasive Mechanical Ventilation (IMV) is considered a life-saving treatment for COVID-19 patients and a huge demand for IMV devices was reported globally. This review aims to provide insight on the initial IMV practices for COVID-19 patients in the initial phase of the pandemic.

**Methods:** Electronic databases (Embase and MEDLINE) were searched for applicable articles using relevant keywords. The references of included articles were hand searched. Articles that reported the use of IMV in adult COVID-19 patients were included in the review. The NIH quality assessment tool for cohort and cross-sectional studies was used to appraise studies.

**Results:** 106 abstracts were identified from the databases search, of which 16 were included. 4 studies were included in the meta-analysis. In total, 9988 patients were included across all studies. The overall cases of COVID-19 requiring IMV ranged from 2–75%. Increased age and pre-existing comorbidities increased the likelihood of IMV requirement. The reported mortality rate in patients receiving IMV ranged between 50–100%. On average, IMV was required and initiated between 10–10.5 days from symptoms onset. When invasively ventilated, COVID-19 patients required IMV for a median of 10–17 days across studies. Little information was provided on ventilatory protocols or management strategies and were inconclusive.

**Conclusion:** In these initial reporting studies for the first month of the pandemic, patients receiving IMV were older and had more pre-existing co-morbidities than those who did not require IMV. The mortality rate was high in COVID-19 patients who received IMV. Studies are needed to evaluate protocols and modalities of IMV to improve outcomes and identify the populations most likely to benefit from IMV.

## Background

In December 2019, an outbreak of pneumonia of unknown cause first emerged in Wuhan, China, and spread rapidly in many regions of that country. Several laboratories identified the causative agent as novel coronavirus (1–3), named as severe acute respiratory syndrome coronavirus-2 (SARS-CoV-2) with the disease termed coronavirus disease 2019 (COVID-19) (4). Due to the highly contagious nature of the disease and the increasing number of countries impacted, COVID-19 was declared a pandemic by the World Health Organization (WHO) on March 11, 2020 (4).

SARS-CoV-2 belongs to the family of single-stranded RNA viruses which includes Middle East Respiratory Syndrome virus (MERS-CoV) and SARS-CoV, which were found to be accountable for previous respiratory syndrome outbreaks (3, 5). Clinical presentation and symptoms of COVID-19 are similar to MERS and SARS; however, the rate of spread is far greater in COVID-19 (6). Furthermore, COVID-19 is the first pandemic disease caused by the coronavirus family (6). As of 22^nd^ of May 2020, the total number of confirmed cases of COVID-19 was 5,263,710 with 337,852 deaths worldwide, and the number is expected to rise (4).

While most people with COVID-19 develop mild or uncomplicated disease, some develop severe illness requiring hospitalisation, with a smaller proportion experiencing respiratory failure requiring Intensive Care Unit (ICU) support (1). In these severe cases, COVID-19 has been causally associated with Acute Respiratory Distress Syndrome (ARDS) requiring ventilatory support including invasive mechanical ventilation (IMV), sepsis, and multiorgan failure, making patient management extremely challenging.

ARDS is characterised by pulmonary infiltrates on imaging such as chest radiographs, a reduction of lung compliance and hypoxemia that is associated with high mortality rates (7). The use of IMV is a crucial intervention for patients developing ARDS (8) but different strategies have been used to mechanically ventilate ARDS patients. To provide an evidence base and unify practice, The ARDS Network (ARDSnet) (a research network that includes the National Heart, Lung, and Blood Institute of the National Institutes of Health (NIH, USA)) was formed to conduct multicentre clinical trials for ARDS treatment (9). The ARDSnet protocol has two approaches in managing ARDS patients requiring mechanical ventilation including either low Positive End-Expiratory Pressure (PEEP) and high Fractional inhaled Oxygen (FiO2) or high PEEP and low FiO2 (8). Currently, there are no guidelines to support ventilatory strategies which are specific to ARDS in COVID-19 patients and recommendations are based on general intensive care management (10) with comorbidity and age effecting mortality rate (11). However, there have been reports of different possible phenotypes in COVID-19 related ARDS that may require IMV strategies that do not follow traditional ARDS IMV protocols(12).

Due to the global outbreak, many health care systems were trying to meet the substantial demand for mechanical ventilators causing a shortage of supplies around the world. The criteria for selection for IMV and protocols used to deliver IMV appear to differ between centres. This systematic review was conducted to describe the utilisation of IMV in patients with COVID-19 at the early stages of this pandemic, specifically to determine if there were patient-driven clinical features or/and IMV technical approaches associated with better outcomes.

## Methods

This review was prepared in accordance to the Preferred Reporting Items for Systematic Reviews and Meta-Analyses (PRISMA) guidelines (13) and the review protocol is registered in the international registry of systematic reviews (PROSPERO) (Registration number CRD42020178262).

Search queries were carried out on the 23^rd^ of April, 2020, using the search strategy shown in (additional file 1) on the following electronic databases: Embase, MEDLINE. http://Clinicaltrails.gov and EudraCT were searched for active trials or published data. Both through scoping searches and discussion with experts, the following search terms were used “COVID-19 OR SARS-COV-2 AND Mechanical ventilation”. The time frame for the search was limited from December 2019 to 23^rd^ of April 2020. Only English language articles were included in the review, but no other limitation was used in the search queries. Search results were imported into EndNote 9.1 (Clarivate Analytics) where duplicates were removed and data was uploaded to Rayyan software(14), a webapp tool used for screening titles and abstracts.

### Eligibility Criteria

Studies were considered for inclusion if they used IMV as part of the treatment of COVID-19 patients. Only patients with confirmed COVID-19 (defined as positive results on real-time reverse transcriptase-polymerase chain reaction (RT-PCR) assay of nasopharyngeal swabs) were included in the review. Studies were considered for exclusion if they were reviews, qualitative studies, and correspondence letters. Studies on individuals under 18 years or who were not treated with IMV were also considered for exclusion.

### Study selection and data extraction

Abstracts were screened blindly and independently by MA and EA using the predefined inclusion and exclusion criteria. Disagreements were resolved by discussion otherwise by MAA. Full-text articles were acquired and imported into EndNote 9.1 by MA and similar abstract screening methodology was used in screening full texts for eligibility. Figure 1 shows studies selection criteria based on PRISMA guidelines.

Data was extracted by NYA and checked by EA for consistency and accuracy using a custom piloted data extraction form developed by MMA and NYA. For each included study, data was extracted on study design, patient characteristics, Intervention and outcomes. Corresponding author were contacted if data were ambiguous or missing.

### Quality Assessment

Quality of included studies was assessed using NIH Quality assessment tool for observational cohort and cross-sectional studies with the quality classified as good, fair and poor. Two independent reviewers assessed quality of included studies (MA and EA). Disagreement was resolved through discussion.

### Data synthesis

Studies were grouped based on disease severity and comorbidities if possible. A narrative synthesis was carried out to assess the proportion of COVID-19 patients requiring invasive mechanical ventilation, when IMV was initiated after symptom onset, and for how long patients received IMV until either clinical improvement or death. Results from studies that provided data on proportion of patients needing IMV were quantitatively synthesized via effects size with random effect meta-analyses using Metaporp (15). Statistical heterogeneity of the included articles was assessed by I^2^ value. Quantitative data was graphed on bar charts and scatterplots to provide graphical representations of the studies.

## Results

### Study selection

Initial searches identified 106 abstracts, of which 7 were duplicates. After abstract screening for eligibility criteria, 18 were included for full text screening. During full text screening, 7 articles were identified that met inclusion criteria and through hand searching of articles references, 9 further articles were identified, making a total of 16 articles to be included in the review. Figure 1 shows the PRISMA flow diagram. http://Clinicaltrials.gov and EudraCT was searched for data and active trials on mechanical ventilation use in COVID-19 patients. There were 902 trials found in http://clincaltrials.gov and 120 trials in EudraCT databases Only one study that was registered on http://clinicaltrials.gov in the Netherlands to retrospectively evaluate IMV modalities and its outcomes on COVID-19 patients (16).

**Figure 1.**
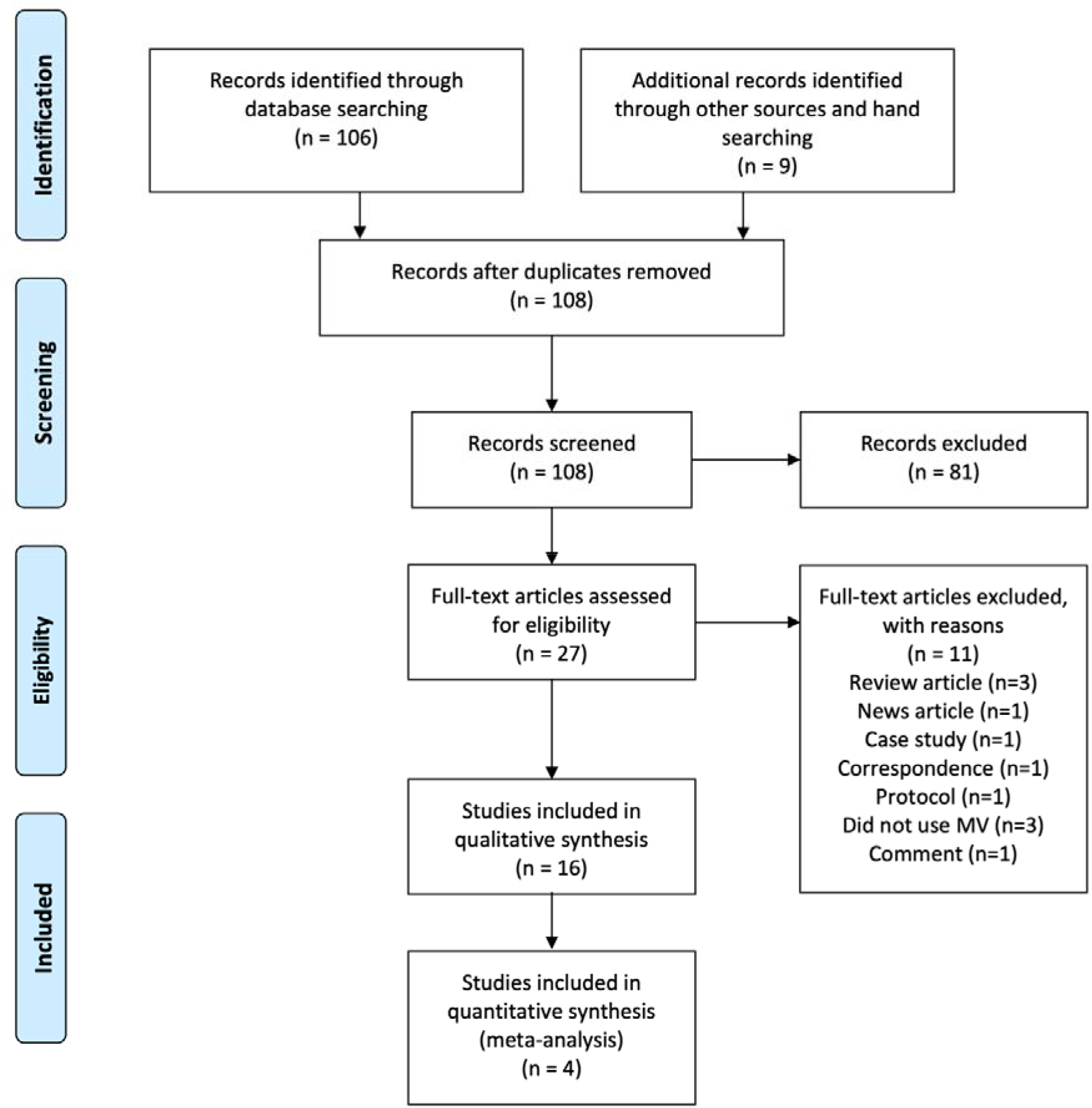
PRISMA flow chart.

### Study Characteristics

Characteristics of the 16 included studies are summarised in table 1. In total, 9988 confirmed COVID-19 patients were included in the selected studies. The sample size of the included studies ranged from 17 to 5700 patients (median 146.5 patients (Interquartile Range (IQR) 49.25 to 254.75)). Of the included studies, all were retrospective apart from one prospective study (17) and most of the studies (12/16) were conducted in China. Of the sixteen included studies, five included only severe cases (defined as patients admitted to ICU or using a stated severity score)(18–22) and eleven included patients consecutively regardless of severity levels (11, 17, 23–31). Although three studies reported the severity of COVID-19 patients, different definitions of severity were used. All studies reported co-morbidities, with hypertension, diabetes mellitus, and cardiovascular diseases the most common, described in table 1. All studies reported the percentage of COVID-19 patients required IMV. However, the baseline characteristics of ventilated versus non-ventilated patients (describing their age, or co-morbidities) were not reported in most of the studies. Furthermore, the criteria for ICU admission was only reported in one study (17) while the criteria for placing patients on IMV was not reported in any study. Definitions of ARDS were reported by nine studies; four used the WHO definition (17, 22, 31, 32) and five used the Berlin definition of ARDS (11, 19, 21, 29). None of the included studies reported whether patients with advanced directives (such as Do Not Resuscitate [DNR]) were included, except one(19).

**Table 1.** characteristics of included studies and reported outcomes.

| Study ID<br>(country) | Groups | Sample Size<br>(Gender) | Age,<br>Mean ±<br>SD or<br>Median<br>(range) | Co-morbidity, N (%) | | | | | | Patients<br>on IMV | Overall<br>Mortality<br>rate | Mortality<br>rate on<br>IMV | Days<br>on<br>IMV | Onset<br>of MV | Reported<br>MV<br>setting<br>(Y/N) |
| --- | --- | --- | --- | --- | --- | --- | --- | --- | --- | --- | --- | --- | --- | --- | --- |
|  |  |  |  | HTN | DM | CVD | CLD | Cancer | Others |  |  |  |  |  |  |
| Liu K. et<br>al., 2020,<br>(China)<sup>a</sup> | Total | 56 | NA | NA | NA | NA | NA | NA | NA | 7<br>(12.5%) | 3 (5.3%) | NR | NR | NR | N |
|  | Young<br>and<br>middle<br>aged | (n=38)<br>Male= 19<br>Female=19 | 47<br>(IQR<br>35.75 -<br>51) | 5(13.16) | 1(2.63) | 0(0) | 0(0) | 0(0) | 1(2.63%) | 3<br>(7.89%) | 2<br>(5.26%) |  |  |  |  |
|  | Elderly | (n=18)<br>Male=12<br>Female=6 | 68<br>(IQR<br>65.25-69.75) | 5(27.78) | 3(16.67) | 2(11.11) | 0(0) | 0(0) | 3<br>(16.67%) | 4<br>(22.22%) | 1(5.56%) |  |  |  |  |
| Mo P. et<br>al., 2020,<br>(China)<sup>b</sup> | Total | 155 | 54<br>(IQR<br>42-66) | 37<br>(23.9) | 15 (9.7) | 15 (9.7) | 8<br>(5.1) | 7 (4.5) | 22 (14.1) | 35<br>(20.6%) | 22<br>(14.2%) | NR | NR | NR | N |
|  | General | (n=70)<br>Male=31<br>Female= 39 | 46<br>(IQR<br>35-56) | 15<br>(21.4) | 3 (4.3) | 0(0) | 0(0) | 2 (2.9) | 4 (5.8) | 0 (0) | NA |  |  |  |  |
|  | Refractory | (n=85)<br>Male= 55<br>Female= 30 | 61<br>(IQR<br>51-70) | 22<br>(25.9) | 12<br>(14.1) | 14<br>(16.5) | 7<br>(8.2) | 5 (5.9) | 18 (11.6) | 35<br>(41.2%) | NA |  |  |  |  |
| Guan et al.,<br>2020<br>(China)<sup>c</sup> | Total | 1099 | 47<br>(IQR<br>35-58) | 165 (15) | 81 (7.4) | 27 (2.5) | 12<br>(1.1) | 10<br>(0.9) | 48 (4.3) | 25<br>(2.3%) | 15<br>(1.4%) | NR | NR | NR | N |
|  | Non-severe | (n=926)<br>Male= 540<br>Female= 386 | 45<br>(IQR<br>34-57) | 124<br>(13.4) | 53 (5.7) | 17 (1.8) | 6<br>(0.6) | 7 (0.8) | 40 (3.6) | 0 (0) | 1 (0.1%) |  |  |  |  |
|  | Severe | (n=173) | 52 | 41 | 28 | 10 (5.8) | 6 | 3 (1.7) | 8 (4.6) | 25 | 14 |  |  |  |  |
|  |  | Male=100<br>Female= 73 | (IQR<br>40-65) | (23.7) | (16.2) |  | (3.5) |  |  | (14.5%) | (8.1%) |  |  |  |  |
| Guo et al.,<br>2020<br>(China)<sup>a</sup> | All | 187 | 58.5 ±<br>14.66 | 61<br>(32.6) | 28 (15) | 21<br>(11.2) | 4<br>(2.1) | 13 (7) | 14 (7.5) | 45 (24%) | 43<br>(22.9%) | NR | NR | NR | N |
|  | Normal<br>TnT | (n=135)<br>Male=57<br>Female=78 | 53.53 ±<br>13.22 | 28<br>(20.7) | 12 (8.9) | 4 (3) | 0 (0) | 7 (5.2) | 1 (0.7) | 14 (10%) | 12<br>(8.8%) |  |  |  |  |
|  | Elevated<br>TnT | (n=52)<br>Male=34<br>Female=18 | 71.4 ±<br>9.43 | 33<br>(63.5) | 16<br>(30.8) | 17<br>(32.7) | 4<br>(7.7) | 6<br>(11.5) | 13 (25) | 31 (60%) | 31 (60%) |  |  |  |  |
| Huang C.<br>et al., 2020<br>(China)<sup>d</sup> | All | 41 | 49<br>(IQR<br>41-58) | 6 (15) | 8 (20) | 6 (15) | 1 (2) | 1 (2) | 1 (2) | 4 (10%) | 6 (15%) | NR | NR | NR | N |
|  | No ICU<br>Care | Male= 18<br>Female= 9 | 49<br>(IQR<br>41-57.5) | 4 (14) | 7 (25) | 3 (11) | 0 (0) | 1 (4) | 1 (4) | 0 | 1 (4%) |  |  |  |  |
|  | ICU Care | Male= 11<br>Female= 2 | 49<br>(IQR<br>41- 61) | 2 (15) | 1 (8) | 3 (23) | 1 (8) | 0 (0) | 0 (0) | 4<br>(30.8%) | 5 (38%) |  |  |  |  |
| Shi S. et<br>al., 2020<br>(China)<sup>a</sup> | All | 416 | 64 (21-95) | 127<br>(30.5) | 60<br>(14.4) | 61<br>(14.5) | 12<br>(2.9) | 9 (2.2) | 47 (11.2) | 32<br>(7.7%) | 57<br>(13.7%) | NR | NR | NR | N |
|  | Without<br>cardiac<br>injury | (n=334)<br>Male= 161<br>Female 173 | 60 (21-90) | 78<br>(23.4) | 40 (12) | 25 (7.5) | 6<br>(1.8) | 2 (0.6) | 27 (8) | 14<br>(4.2%) | 15<br>(4.5%) |  |  |  |  |
|  | With<br>cardiac<br>injury | (n=82)<br>Male= 44<br>Female= 38 | 74 (34-95) | 49<br>(59.8) | 20<br>(24.4) | 36<br>(43.9) | 6<br>(7.3) | 7 (8.5) | 20 (24) | 18 (22%) | 42<br>(51.2%) |  |  |  |  |
| Wang et<br>al., 2020<br>(China)<sup>e</sup> | Total | 17<br>Male=7<br>Female=10 | 65<br>(IQR<br>56-75) | 3 (18%) | 3 (18%) | 3 (18%) | 0 (0) | 0 (0) | 0 (0) | 2<br>(11.7%) | NA | NR | NR | NR | N |
| Yang et al., | Total | 52 | 59.7 | 0 (0) | 9 (17) | 5 (10) | 4 (8) | 2 (4) | 9 (17) | 22 (42%) | 32 | 19 (86%) | NR | NR | N |
| 2020<br>(China)<sup>f</sup> |  |  | ±13.3 |  |  |  |  |  |  |  | (61.5%) |  |  |  |  |
|  | Survivors | (n=20)<br>Male= 14<br>Female= 6 | 51.9 ±<br>12.9 | 0 (0) | 2 (10) | 2 (10) | 2 (10) | 1 (5) | 0 (0) | 3 (15%) |  |  |  |  |  |
|  | Non-survivors | (n=32)<br>Male=21<br>Female= 11 | 64.6<br>±11.2 | 0 (0) | 7 (22) | 3 (9) | 2 (6) | 1 (3) | 9 (28) | 19 (59%) |  |  |  |  |  |
| Grasselli et<br>al., 2020<br>(Italy)<sup>f</sup> | Total<br>including<br>all ages | 1591<br>Male=1304<br>Female= 287 | 63<br>(IQR<br>56-70) | 509 (49) | 180<br>(17%) | 223<br>(21%) | 42<br>(4%) | 81 (8) | 457<br>(43.8) | 1150<br>(72.3%) | 405<br>(26%) | NR | NR | NR | Y |
|  | 21-40 | Male=44<br>Female=12 | 34<br>(IQR<br>31-38) | 4 (11%) | 1 (3%) | 1 (3%) | 1<br>(3%) | 0 | 8 (22.8) | 37 (66%) | 4 (7%) |  |  |  |  |
|  | 41-50 | Male=119<br>Female=24 | 47<br>(IQR<br>44-49) | 21<br>(26%) | 4 (5%) | 4 (5%) | 0 (0) | 2 (2) | 15 (18.2) | 87<br>(60.8%) | 16 (11%) |  |  |  |  |
|  | 51-60 | Male=355<br>Female=72 | 56<br>(IQR<br>54-59) | 121<br>(44%) | 40<br>(15%) | 43<br>(16%) | 8<br>(3%) | 10 (4) | 97 (35) | 315<br>(73.8%) | 63 (15%) |  |  |  |  |
|  | 61-70 | Male=484<br>Female=114 | 65<br>(IQR<br>63-68) | 195<br>(51%) | 86<br>(23%) | 87<br>(23%) | 12<br>(3%) | 33 (9) | 199 (52) | 449<br>(75.1%) | 174<br>(29%) |  |  |  |  |
|  | 71-80 | Male=279<br>Female=62 | 74<br>(IQR<br>72-76) | 156<br>(62%) | 46<br>(18%) | 81<br>(32%) | 20<br>(8%) | 33<br>(19) | 126 (50) | 246<br>(72.1%) | 136<br>(40%) |  |  |  |  |
|  | 81-90 | Male=19<br>Female=2 | 83<br>(IQR<br>81-84) | 12<br>(75%) | 3 (19%) | 6 (38%) | 1<br>(6%) | 3 (19) | 10 | 14<br>(66.6%) | 11 (52%) |  |  |  |  |
|  | 91-100 | Male=1 | 91 | 0 (0) | 0 (0) | 1<br>(100%) | 0 (0) | 0 (0) | 0 (0) | 0 (0) | 1 (100%) |  |  |  |  |
| Bhatraju et<br>al., 2020 | Total with<br>no groups | 24<br>Male=15 | 64 ±18 | 0 (0) | 14 (58) | 0 (0) | 4 (16) | 0 (0) | 17 (70) | 18 (75%) | 12 (50%) | NR | 10<br>(IQR) | NR | Y |
| (USA)<sup>f</sup> |  | Female=9 |  |  |  |  |  |  |  |  |  |  | 7-12) |  |  |
| Arentz et al., 2020 (USA)<sup>f</sup> | Only critical Patients | 21<br>Male= 52%<br>Female= 48% | 70 (43-92) | 0 (0) | 7 (33.3) | 9 (42.9) | 9 (42.9) | 0 (0) | 10 (47.6) | 15 (71%) | 11 (52.4%) | NR | NR | NR | N |
| Wu C. et al., 2020 (China)<sup>a</sup> | All | 201 | 51 (IQR 43-60) | 39 (19.4) | 22 (10.9) | 8 (4) | 5 (2.5) | 1 (0.5) | 18 (9) | 6 (3%) | 44 (21.9%) | 6 (100%) | NR | NR | N |
|  | Without ARDS | (n=117)<br>Male=68<br>Female=49 | 48 (IQR 40-54) | 16 (13.7) | 6 (5.1) | 3 (2.6) | NA | NA | NA | 0 (0) | 0 (0) |  |  |  |  |
|  | With ARDS | (n=84)<br>Male=60<br>Female=24 | 58.5 (IQR 50-69) | 23 (27.4) | 16 (19) | 5 (6) | NA | NA | NA | 6 (7.2%) | 44 (52.3%) |  |  |  |  |
| Chen et al., 2020 (China)<sup>a</sup> | All severities | 99<br>Male= 67<br>Female= 32 | 55.5 ± 13.1 | 40 (40%) | 0 (0) | 0 (0) | 1 (1%) | 1 (1%) | 25 (25) | 4 (4%) | 11 (11%) | NR | NR | NR | Y |
| Zhou et al., 2020 (China)<sup>a</sup> | Total | 191 | 56 (46-67) | 58 (30) | 36 (19) | 15 (8) | 6 (3) | 2 (1) | 24 (12.5) | 32 (17%~~%~~) | 54 (28.2%) | 31 (97%) | NR | 14.5 days (12.0-19.0) | N |
|  | Survivors | (n=137)<br>Male= 81<br>Female= 54 | 52 (45-58) | 32 (23) | 19 (14) | 13 (24) | 2 (1) | 0 (0) | 11 (8) | 1 (1%) |  |  |  |  |  |
|  | Non-survivors | (n=54)<br>Male=38<br>Female= 16 | 69 (63-76) | 26 (48) | 17 (31) | 2 (1) | 4 (7) | 2 (1) | 13 (24) | 31 (57%) |  |  |  |  |  |
| Wang et al., 2020 (China)<sup>d</sup> | Total | 138 | 56 (IQR 42-68) | 43 (31.2) | 14 (10.1) | 20 (14.5) | 4 (2.9) | 10 (7.2) | 17 (12.3) | 17 (12.32%) | 6 (4.3%) | NR | NR | NR | N |
|  | Non-ICU | (n=102)<br>Male= 53<br>Female=51 | 51 (IQR 37-62) | 22 (21.6) | 6 (5.9) | 11 (10.8) | 1 (1) | 6 (5.9) | 9 (8.8) | 0 |  |  |  |  |  |
|  | ICU | (n=36)<br>Male=22<br>Female=14 | 66<br>(IQR<br>57-78) | 21<br>(58.3) | 8 (22.2) | 9 (25) | 3<br>(8.3) | 4<br>(11.1) | 8 (22.2) | 17 (41.7) |  |  |  |  |  |
| Richardson<br>S et al.<br>(USA)<sup>a</sup> | All<br>Patients,<br>no groups | 5700<br>Male=3437<br>Female=2263 | 63<br>(IQR<br>52-75)<br>(range<br>0-107) | 3026<br>(56.6) | 1808<br>(33.8) | 966<br>(16.9) | 920<br>(16.1) | 320<br>(6) | 3110<br>(54.5) | 1151<br>(20.2) | 553 (21) | 282<br>(88.1%) | NR | NR | N |

<sup>a</sup> = study included patients on both general and critical care.
<sup>b</sup> = despite the reported severities, they grouped patients into those with symptoms resolution, improvements in radiological abnormalities, hospital stay $\leq 10$ days, and the maintenance of body temperature with the use of antipyretics (general or responders) and those without improvement (refractory).
<sup>c</sup> = severity was assessed at the time of admission using the ATS for community-acquired pneumonia (severe vs. non-severe).
<sup>d</sup> = Critical patients were defined by the need for ICU care.
<sup>e</sup> = Severe patients are defined by the need for HFNC, NIV or IMV to improve oxygenation.
<sup>f</sup> = Critical patients defined by the admission to ICU.
Elderly= $\geq 60$ years old, young and middle aged= $< 60$ years old, IQR= Interquartile range, ICU= Intensive Care Unit, ATS= American Thoracic Society, TnT= troponin T, N= No, Y= Yes, HFNC= high flow nasal cannula, NIV= Non-invasive mechanical ventilation, IMV= invasive mechanical ventilation, HTN=Hypertension, DM= Diabetes Mellitus, CVD=Cardiovascular Disease, CLD=Chronic Lung Disease.

### Quality assessment

As the studies were cohort studies and case-series, all included articles were assessed using the pre-specified quality assessment tool, the NIH quality assessment tool for cohort/cross-sectional studies. Domains 5, 9, 10, 12 and 13 of the NIH quality assessment tool were not assessed as they relate to power calculation, exposure measure overtime, blinding of outcome assessment, and loss to follow-up over time, and these were not applicable to the included studies. The summary graph of the included studies can be found in additional file 2. Only 3/16 studies were of a fair quality (18, 26, 27), otherwise all studies 13/16 were of good quality based on the quality assessment tool.

### Result of individual studies

#### Proportion of COVID-19 patients receiving IMV

The proportion of COVID-19 patients receiving invasive mechanical ventilation are reported based on their inclusion criteria, with studies that included only severely unwell patients grouped together and studies that included patients consecutively regardless of severity status reviewed in a separate grouping.

#### Severe or Critical cases

Five studies assessed the characteristics and outcomes of severe or critically ill patients with COVID-19 and reported the proportion of patients requiring IMV. Grasselli et al. reported that 72.3% (1150/1591) of the patients admitted to ICU required IMV. These authors also grouped the patients by age range with 73.8% (315/427) of patients aged 51–60 years requiring IMV; 75.1% (449/598) of patients aged 61–70 years and 72.1% (246/341) of patients aged 71–80 years requiring IMV(20). Criteria used to assess the need for IMV were not reported. Bhatraju et al. reported that IMV was initiated in 79.2% (19/24) of critically ill COVID-19 patients with no description of the criteria used to assess the need for IMV(19). Arentz et al. included only critically ill patients in their study (defined as requiring ICU support) and here 71% (15/21) of COVID-19 patients received IMV(18). These authors also stated that ARDS was observed in all patients requiring IMV but gave no further details of the decision process for IMV use.

Yang et al. also included critically ill patients and reported that IMV was initiated in 42% (22/52) of patients(33). When the patients were grouped into survivors and non-survivors, 15% (3/20) of the survivors required IMV, while 59% (19/32) of those who had died required IMV but they did not explicitly relate this to disease severity. Wang et al. assessed the use of High Flow Nasal Cannula (HFNC) in severe COVID-19 patients and reported that 41.1% (7/17) failed HFNC and were placed on NIV as rescue therapy(21). Of the seven patients on NIV, two failed and required IMV. Therefore, in this small study, 11.7% of the patients placed initially on HFNC (2/17) required IMV. Figure 2 shows the percentages of severe or critical patients that required IMV and the weighted mean across all studies.

**Figure 2.**
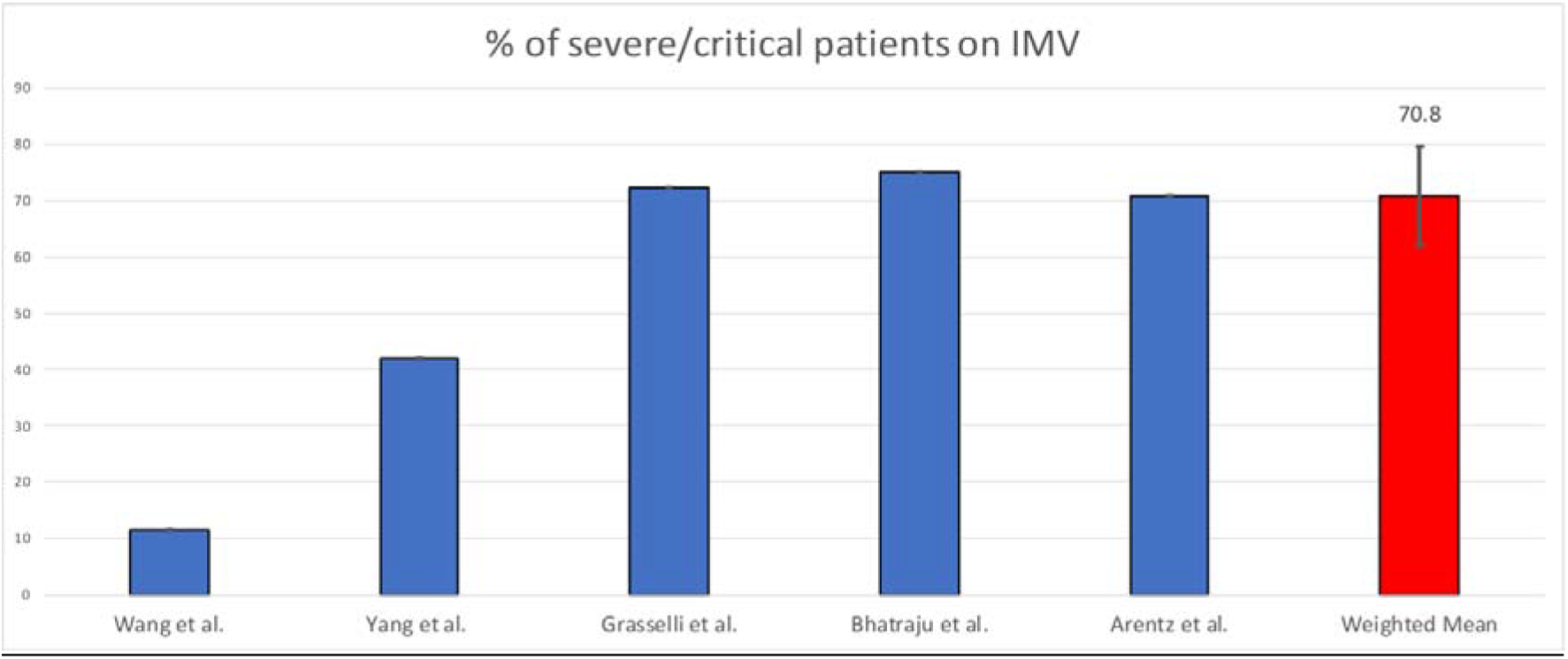
Percentage of severe or critical COVID-19 patient requiring IMV.

#### Consecutive sampling

The proportion of COVID-19 patients required IMV was also reported in eleven studies where patients were sampled consecutively, and severity of the disease was not specified at recruitment. Six of these studies aimed to assess the clinical characteristics of patients with COVID-19. Liu et al. included 56 patients and reported the percentage of patients required IMV in two groups, separated by age(26). In the elderly (defined as ≥ 60 years old), 22.2% (4/18) required IMV while in young to middle-aged patients (aged < 60 years old), 7.9% (3/38) required IMV. Mo et al. included 155 patients with different severities of COVID-19 and reported that 20.6% (35/155) of the patients required IMV(27). Furthermore, this study divided patients into two general groups at day 10 of hospitalization (defined into “responders” which included those with symptoms resolution, improvements in radiological abnormalities, hospital stay ≤ 10 days, and the maintenance of body temperature with the use of antipyretics and “refractory” where these parameters had not been met). In refractory COVID-19 patients, 41.2% (35/85) required IMV while in the responding patients, IMV was not required.

A retrospective cohort study by Guan et al. also evaluated the clinical characteristics of COVID-19 in 1099 patients, and they reported that 2.3% (25/1099) required IMV. (24) When patients were grouped into severe and non-severe based on the ATS Community Acquired Pneumonia guidelines, all those who required IMV were from the severe patients’ group. Among the severe patients, 14.5% (25/173) required IMV but the study did not report their demographics separately. Huang et al. conducted a prospective cohort study to assess the clinical feature of COVID-19 patients and reported that out of the 41 patients included, 13 required admission to the ICU (29). Of these, NIV was used in 61.5% (8/13), one (7.7%) patient required oxygen provided by nasal cannula and IMV was initiated in 30.8% (4/13). Of those receiving IMV, two had refractory hypoxemia and received Extracorporeal Membrane Oxygenation (ECMO). Wang et al. included 138 COVID-19 patients in their retrospective study and reported that 36 of the patients required ICU care (30). They also reported that among these ICU patients, 47.2% (17/36) required IMV, 41.7% (15/36) required NIV and 11.1% (4/36) required oxygen inhalation. Chen et al. did not specify the severity of the included COVID-19 patients but reported that 4.0% (4/99) required IMV(25). A large case series by Richardson S. et al., which described the clinical features and outcomes of hospitalised patients with COVID-19, reported that 20.2% (1151/5700) required IMV(28).

A retrospective study by Zhou et al. aimed to assess the clinical course and risk factors for mortality of adult COVID-19 patients and reported that 17% (32/191) of the patients required IMV(11). When patients were grouped into survivors and non-survivors, 57% (31/54) of the non-survivor patients required IMV. Wu C. et al. aimed to assess the risk factors associated with ARDS and death in COVID-19 patients(31). The authors reported that 84/201 patients developed ARDS and of those, 7.2% (6/84) required IMV and of those, one patient (2.3%) received ECMO. The authors also reported that NIV and regular nasal cannula were used in 72.6% (61/84) and 20.2% (17/84) of patients with ARDS, respectively with ARDS defined using the WHO interim definitions(34).

Two retrospective studies assessing the association of cardiac injury with death in patients with COVID-19 also reported the percentage of patients required IMV. Guo et al. included 187 COVID-19 patients and grouped them into two groups by their serum levels of troponin T (TnT) to determine their risk of cardiac injury (normal and elevated (defined by above the 99^th^ percentile upper reference limit))(25). In total, 25% (45/187) required IMV, and in patients with elevated TnT, 60% (31/52) required IMV while in patients with normal TnT, only 10% (14/135) required IMV. In a cohort study by Shi et al, 416 patients with COVID-19 were grouped into those with cardiac injury and without cardiac injury (defined as blood levels of cardiac biomarkers (hs-TN1) above the 99^th^ percentile upper reference limit)(29). 7.7% (32/416) of the total cohort required IMV. In patients with cardiac injury, 22% (18/82) required IMV but in those without cardiac injury, 4.2% (14/334) required IMV. The percentage of COVID-19 patients required IMV of studies included patients consecutively regardless of severity status are shown in figure 3.

**Figure 3.**
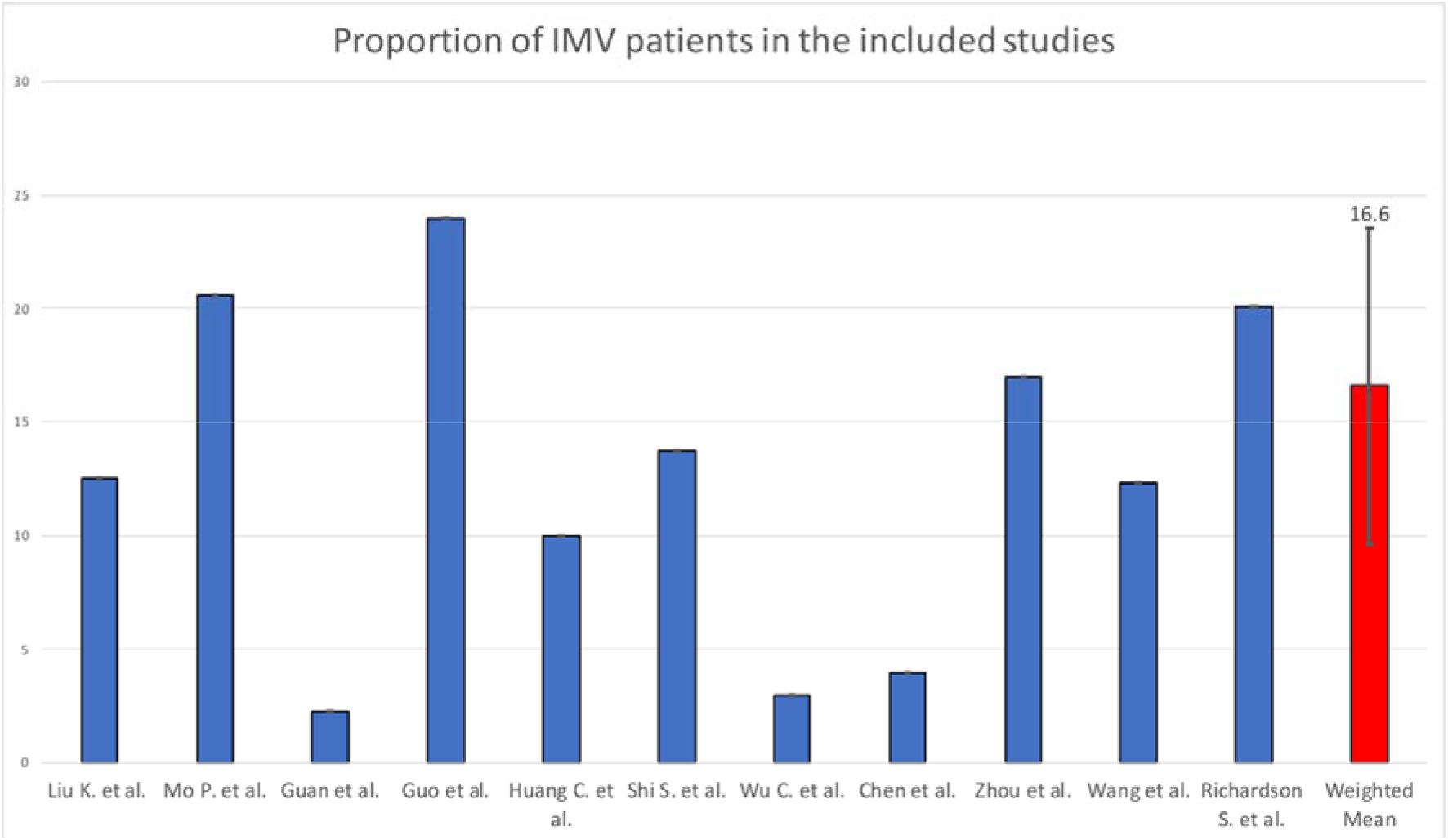
Percentage of the patients that required IMV from studies of consecutive sampling and weighted mean.

#### Mortality rate in patients who received IMV

Five studies reported the mortality rate of patients receiving IMV rather than overall mortality. Wu C. et al. reported a 100% mortality rate (n = 6) for mechanically ventilated patients with ARDS in all the IMV patients(31). Zhou et al. reported a high mortality rate of 97% for the 32 IMV patients, with only 1 patient who survived(11). Yang et al. reported a mortality rate of 86% of 22 IMV patients(22). Richardson et al. reported a mortality rate of 88% of the (320/1151) ventilated patients with a reported outcome in the study(28). Bhatraju et al. reported a mortality of 50% of 18 IMV patients, but 5 patients were still on IMV at end of the study period and only 4/18 (22%) survived to discharge. Figure 4 illustrates mortality rate and survival rate of patient on IMV(19). Overall disease severity and mortality were associated with the presence of comorbidities, with hypertension, diabetes, and cardiovascular diseases the most commonly reported.

**Figure 4.**
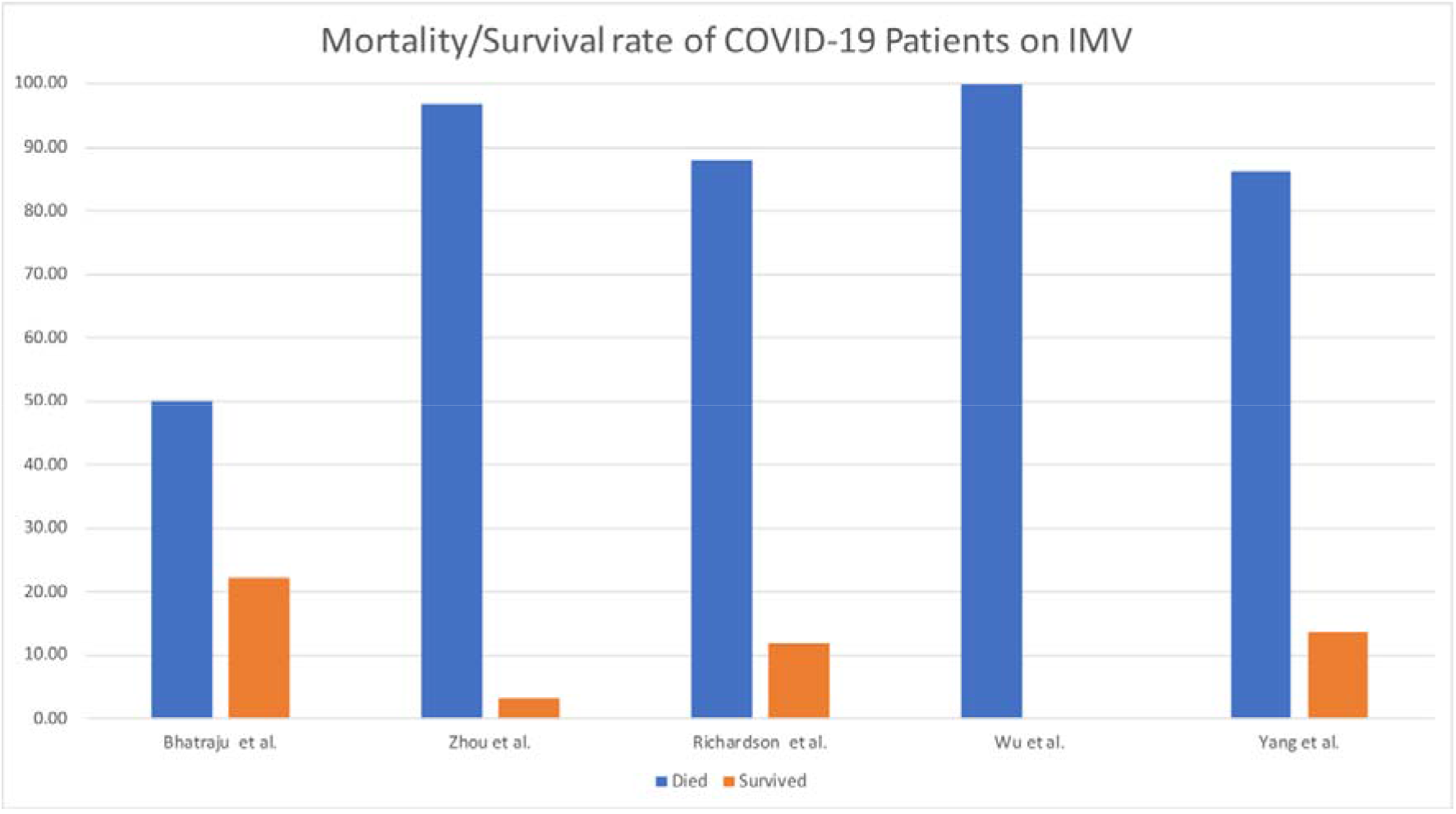
Mortality and survival rate of COVID-10 patients on IMV.

### Ventilator utilization

#### Duration of IMV

Only 2 studies reported the duration of IMV for COVID-19 patients (see additional file 3). Bhatraju et al. reported a median duration for IMV 10 days (IQR 7–12) (19). In a sub-analysis, they found that surviving patients had a slightly higher duration of IMV than the overall sample with a median of 11 days (IQR 7–12). Chen et al. reported a median duration of 17 days (IQR 12–19) for IMV, but they had a smaller IMV population of 4 patients(23).

#### Initiation of IMV in the course of the illness

Only 2 studies reported when IMV was instigated compared to onset of symptoms. Huang. C. et al. reported that in the 4 patients that required MV, the onset of MV was at a median of 10.5 days (IQR 7.0–14.0) from the onset of symptoms(17). Zhou et al. reported a similar median of MV onset of 10.0 days (IQR 5.0–12.5) in a larger population of 32 patients(11). See figure 5.

**Figure 5.**
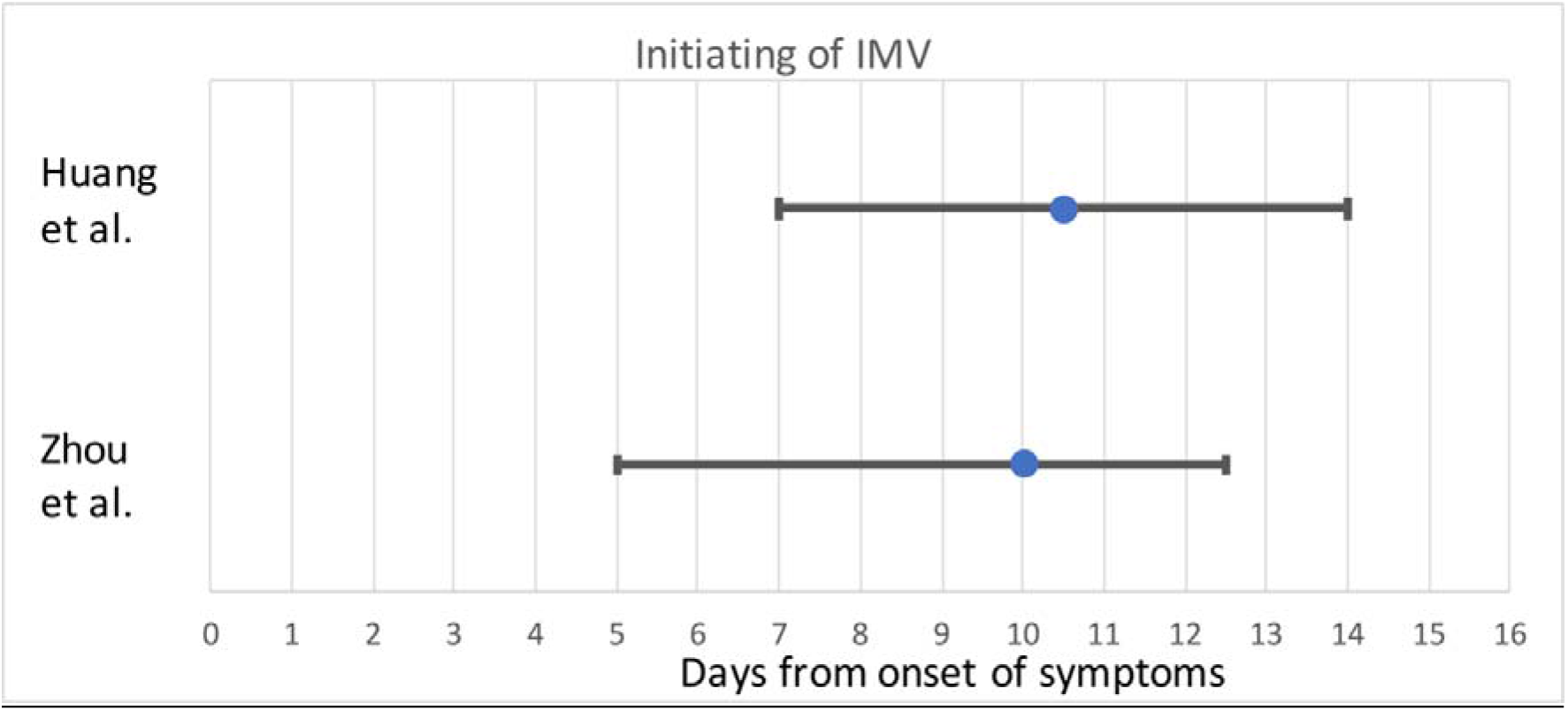
The initiation of IMV in COVID-19 patients.

#### Ventilator setting

None of the included studies were designed to assess the effectiveness of IMV modalities in COVID-19 patients, but 3 studies reported some IMV settings. Grasselli et al. reported IMV settings for specific age groups(20). The overall median PEEP setting was 14cm H_2_O (IQR 12–16) and this was used for most age groups apart from the oldest (81 – 90years) where a median PEEP of 12cmH_2_O (IQR 8–15) was reported. Bhatraju et al. reported FiO2, but did not discuss other ventilatory settings other than IMV readings of plateau pressure, driving pressure, and compliance over the first three days of MV(19). On the first day of IMV, the median FiO2 was set at 0.9 and readings included a median plateau pressure of 25 cmH_2_O (IQR 20–28) and median compliance 29 mL/cmH_2_O (IQR 25–36). On the second day, there was a titration of FiO2 (0.7) as lung mechanics were improving. More improvement was reported on the third day of MV allowing more titrations of FiO2 to 0.6. Chen et al. reported general settings used on the 4 ventilated patients as using P-SIMV with a PEEP of 6–12 cmH2O and FiO2 between 35–100%(23).

### Data synthesis

The overall published studies included in this systematic review suggested that only a small proportion of COVID-19 patients required IMV. But in severe cases the overall effect size of IMV is 65% (95% CI: 49–80) I^2^ 84.37%. Figure 6 shows pooled data of the effect size on a forest plot.

**Figure 6.**
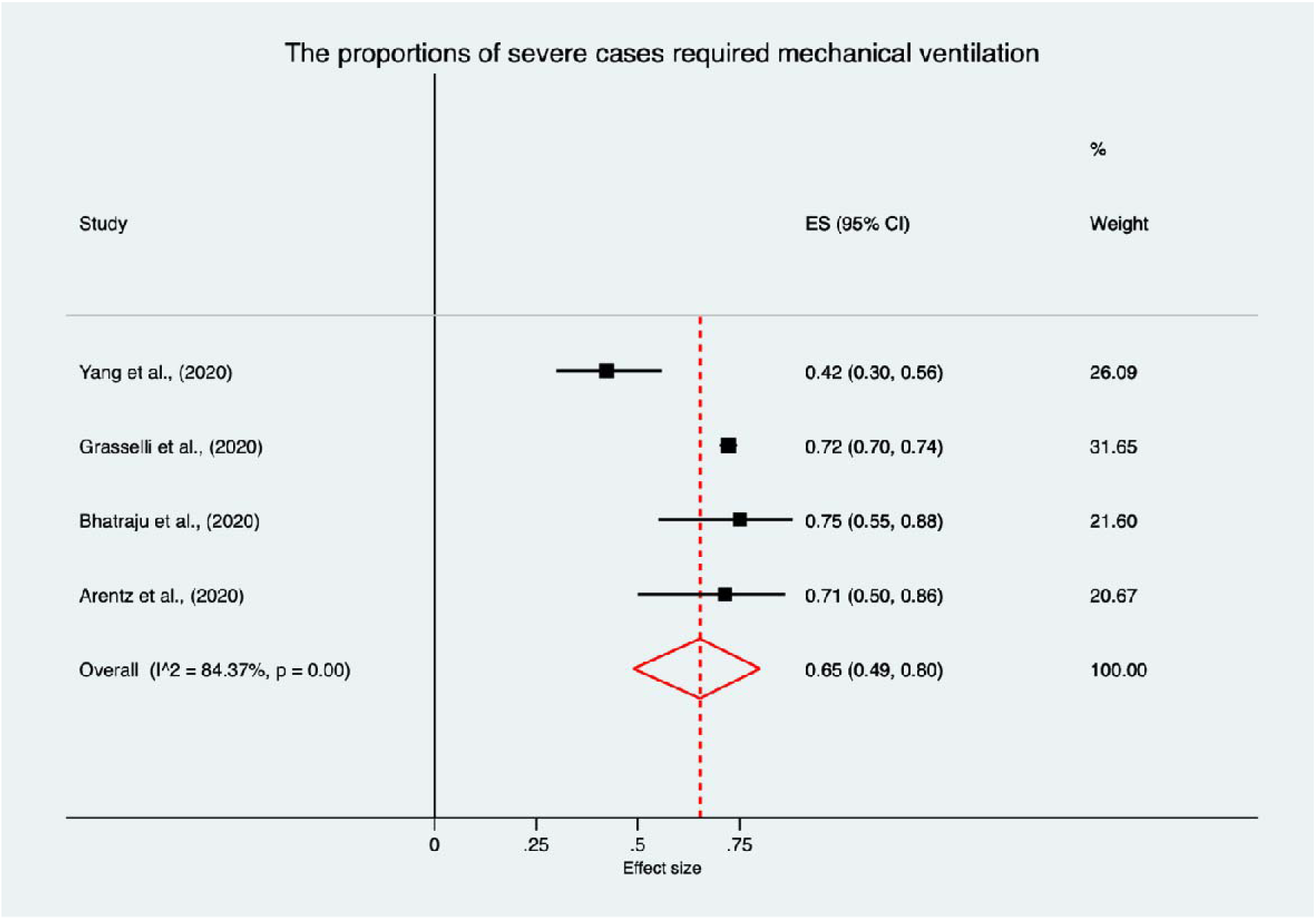
Forest plot of the proportions of severe cases requiring IMV.

## Discussion

To the best of our knowledge, this is the first systematic review that assesses the utilization of IMV in COVID-19 patients. IMV practices are likely to change as our experience of this condition increases (including not only who receives IMV but also the protocols used). Objectively summarising the studies published within the first month of the pandemic provides an important benchmark, by which to compare changing practices and outcomes for patients.

The use of IMV is critical in treating ARDS and many studies and protocols have been developed in order to manage it appropriately (8). Current guidelines and studies regarding the use of IMV for COVID-19 patients suggest clinicians are managing patients with COVID-19 related ARDS using similar ventilatory strategies to those adopted for other forms of ARDS (35). While that may be an appropriate approach to manage ARDS, it may not be the optimal management strategy in COVID-19. Unfortunately, no reported studies have compared different ventilatory strategies in COVID-19. Only 1 trial is currently registered in http://clinicaltrials.gov and none in EudraCT are studying the modalities of IMV in COVID-19 patients. The trial (PRoVENT-COVID) is a multicentre clinical trial that is designed to compare different ventilatory management strategies to determine how ventilator settings effect the duration of IMV support in COVID-19 (16).

In general, most of the included studies in this review (12/16) are from China, (3/16) the United States (US) and one (1/16) was from Italy. Based on the quality assessment tool used in this review, the methodological quality of included studies was good except for three studies, which were of fair quality.

In studies which only included patients on critical care, the proportion of COVID-19 patients who received IMV ranged between 11–88%. One study suggested that non-invasive respiratory support devices such as HFNC was associated with lower IMV usage(21), but the study was small. In studies which included consecutive COVID-19 patients (with patients on general wards and critical care units), the proportion of IMV use was between 2–20%, with a weighted total of 16%. Mortality in those receiving IMV was high. However, the characteristics of patients that received IMV were not reported in all studies and the protocols for assessing when IMV use was required and ventilatory strategies or settings were not discussed.

There were differences in the prevalence of IMV use across studies and between countries. For example, IMV use was lower in China in severely unwell patients admitted to ICU, especially in younger patients(11, 22) ranging from (14–41%)(17, 32, 36) compared to studies in Italy and the US, where 71–75% of patients received IMV(18–20). The reasons for this are unclear, but could reflect differences in the course of the illness (potentially patients in Italy and USA may have experienced a more aggressive form of COVID-19), or patient factors including the influence of age, ethnicity, co-morbidity and frailty) or differences in care escalation decisions.

In COVID-19 patients on critical care units, an evidence-based protocol IMV would be beneficial, especially while there are no available, specific treatments for the virus. It is still unclear who would benefit most from IMV, and there is a need to characterise this, considering demographics, co-morbidities and the onset of symptoms. Strategies to delay or minimise IMV use should be considered and the use of HFNC has been found helpful in a small sample of severe patients and warrants further study(21). Other non-invasive ventilatory support methods also appear to be helpful(37), but there are concerns of nosocomial transmission of infection to health care workers(38). This also requires further study. In ARDS patients, nursing patients in the prone position has been found helpful in improving gas exchange and in lowering mortality (39). The use of prone position in COVID-19 patients is being studied to determine whether this improves outcomes or reduces the duration of IMV(40–42).

In these early studies of COVID-19 patients, IMV was associated with low survival rate (0–22%). Nevertheless, overall mortality was linked to demographic factors such as male sex and age as well as co-morbidities including hypertension, diabetes and cardiovascular disease (CVD). In keeping with this, studies assessing cardiac impairment in COVID-19 have reported higher mortality and use of IMV(25, 29).

Only two studies reported the duration of IMV in COVID-19, with Bhatraju et al. reporting IMV duration of 10 days (IQR 7–12)(19) and Chen et al. reporting a median duration of IMV of 17 days (IQR 12–19)) (23). This variation may reflect the size of the studies and the population included. Bhatraju et al. only included patients on critical care while in Chen et al. included both patients on general care and critical care. Furthermore, Chen et al. had only 4 patients required IMV while in Bhatraju et al. IMV was received by 18 patients. Data from two studies reporting the initiation of IMV in the course of the illness showed that IMV was usually required approximately 10 days after the onset of symptoms.

This systematic review was limited by the small number of studies included, and they were mostly conducted in China and thus may not represent the wider COVID-19 experience. The evidence-base is continually expanding, but by summarising the initial experience of the clinical community, this review serves two main purposes. First to highlight gaps in our knowledge. The lack of detail about ventilatory protocols and patient demographics limits our understanding on which patients may have better outcomes with IMV and how they should be treated, and further clarification is required. Second, it provides a benchmark from which subsequent reviews can compare changes in practice and their impact on outcome.

## Conclusion

Invasive mechanical ventilation is a life-saving treatment in COVID-19 patients, yet the mortality rate remains very high. Modalities of IMV need to be studied in order to develop appropriate protocols and manage patients effectively. The use of non-invasive ventilatory support may reduce IMV usage, but further studies are needed to evaluate the risk and benefit of using such modalities. Where IMV is a limited resource, patient factors such as co-morbidities, may help identify those who are most likely to benefit from IMV, but further studies are required.

## Data Availability

All data generated or analysed during this review are obtained from the included peer-reviewed published articles.

## Authors’ contributions

MAA conceived, planned, and analysed the data, and major contributor in writing the manuscript. NYA conceived, planned, and analysed the data, and major contributor in writing the manuscript. MA performed abstract screening, full-text screening and quality assessment. EA performed abstract screening, full-text screening and quality assessment. ZA planned and performed searching strategy and data synthesis. FA contributed in the design and analysis. ES planned and designed the review, and major contributor in writing the manuscript and interpretations. DP planned and designed the review, and major contributor in writing the manuscript and interpretations. All authors have read and approved the manuscript.

## Notes

### Competing Interest Statement

The authors have declared no competing interest.

### Clinical Protocols

https://www.crd.york.ac.uk/prospero/display_record.php?RecordID=178262

### Funding Statement

no external funding was received

## References

1. Lu R, Zhao X, Li J, Niu P, Yang B, Wu H, et al. Genomic characterisation and epidemiology of 2019 novel coronavirus: implications for virus origins and receptor binding. The Lancet. 2020;395(10224):565–74.

2. Zhou P, Yang XL, Wang XG, Hu B, Zhang L, Zhang W, et al. A pneumonia outbreak associated with a new coronavirus of probable bat origin. Nature. 2020;579(7798):270–3.

3. Zhu N, Zhang D, Wang W, Li X, Yang B, Song J, et al. A Novel Coronavirus from Patients with Pneumonia in China, 2019. New England Journal of Medicine. 2020;382(8):727–33.

4. WHO. Coronavirus disease (COVID-2019) Situation Report 2020 [Available from: https://www.who.int/emergencies/diseases/novel-coronavirus-2019/events-as-they-happen..

5. Su S, Wong G, Shi W, Liu J, Lai ACK, Zhou J, et al. Epidemiology, Genetic Recombination, and Pathogenesis of Coronaviruses. Trends Microbiol. 2016;24(6):490–502.

6. Peeri NC, Shrestha N, Rahman MS, Zaki R, Tan Z, Bibi S, et al. The SARS, MERS and novel coronavirus (COVID-19) epidemics, the newest and biggest global health threats: what lessons have we learned? Int J Epidemiol. 2020.

7. Slutsky AS, Ranieri VM. Mechanical ventilation: lessons from the ARDSNet trial. Respiratory research. 2000;1(2):73–7.

8. Higher versus Lower Positive End-Expiratory Pressures in Patients with the Acute Respiratory Distress Syndrome. New England Journal of Medicine. 2004;351(4):327–36.

9. Network ARDS. ARDSnet 2020 [Available from: http://www.ardsnet.org/.

10. Phua J, Weng L, Ling L, Egi M, Lim C-M, Divatia JV, et al. Intensive care management of coronavirus disease 2019 (COVID-19): challenges and recommendations. The Lancet Respiratory Medicine.

11. Zhou F, Yu T, Du R, Fan G, Liu Y, Liu Z, et al. Clinical course and risk factors for mortality of adult inpatients with COVID-19 in Wuhan, China: a retrospective cohort study. The Lancet. 2020;395(10229):1054–62.

12. Marini JJ, Gattinoni L. Management of COVID-19 Respiratory Distress. JAMA. 2020.

13. Liberati A, Altman DG, Tetzlaff J, Mulrow C, Gotzsche PC, Ioannidis JP, et al. The PRISMA statement for reporting systematic reviews and meta-analyses of studies that evaluate healthcare interventions: explanation and elaboration. BMJ. 2009;339:b2700.

14. Ouzzani M, Hammady H, Fedorowicz Z, Elmagarmid A. Rayyan-a web and mobile app for systematic reviews. Syst Rev. 2016;5(1):210.

15. Nyaga VN, Arbyn M, Aerts M. Metaprop: a Stata command to perform meta-analysis of binomial data. Archives of Public Health. 2014;72(1):39.

16. http://clinicaltrials.gov. PRactice of VENTilation in COVID-19 Patients (PRoVENT-COVID) –Full Text View – http://ClinicalTrials.gov.

17. Huang C, Wang Y, Li X, Ren L, Zhao J, Hu Y, et al. Clinical features of patients infected with 2019 novel coronavirus in Wuhan, China. The Lancet. 2020;395(10223):497–506.

18. Arentz M, Yim E, Klaff L, Lokhandwala S, Riedo FX, Chong M, et al. Characteristics and Outcomes of 21 Critically Ill Patients With COVID-19 in Washington State. JAMA. 2020.

19. Bhatraju PK, Ghassemieh BJ, Nichols M, Kim R, Jerome KR, Nalla AK, et al. Covid-19 in Critically Ill Patients in the Seattle Region — Case Series. New England Journal of Medicine. 2020.

20. Grasselli G, Zangrillo A, Zanella A, Antonelli M, Cabrini L, Castelli A, et al. Baseline Characteristics and Outcomes of 1591 Patients Infected With SARS-CoV-2 Admitted to ICUs of the Lombardy Region, Italy. JAMA. 2020.

21. Wang K, Zhao W, Li J, Shu W, Duan J. The experience of high-flow nasal cannula in hospitalized patients with 2019 novel coronavirus-infected pneumonia in two hospitals of Chongqing, China. Annals of Intensive Care. 2020;10(1):37.

22. Yang X, Yu Y, Xu J, Shu H, Xia J, Liu H, et al. Clinical course and outcomes of critically ill patients with SARS-CoV-2 pneumonia in Wuhan, China: a single-centered, retrospective, observational study. The Lancet Respiratory Medicine. 2020.

23. Chen N, Zhou M, Dong X, Qu J, Gong F, Han Y, et al. Epidemiological and clinical characteristics of 99 cases of 2019 novel coronavirus pneumonia in Wuhan, China: a descriptive study. The Lancet. 2020;395(10223):507–13.

24. Guan W-j, Ni Z-y, Hu Y, Liang W-h, Ou C-q, He J-x, et al. Clinical Characteristics of Coronavirus Disease 2019 in China. New England Journal of Medicine. 2020.

25. Guo T, Fan Y, Chen M, Wu X, Zhang L, He T, et al. Cardiovascular Implications of Fatal Outcomes of Patients With Coronavirus Disease 2019 (COVID-19). JAMA Cardiology. 2020.

26. Liu K, Chen Y, Lin R, Han K. Clinical features of COVID-19 in elderly patients: A comparison with young and middle-aged patients. J Infect. 2020.

27. Mo P, Xing Y, Xiao Y, Deng L, Zhao Q, Wang H, et al. Clinical characteristics of refractory COVID-19 pneumonia in Wuhan, China. Clinical Infectious Diseases. 2020.

28. Richardson S, Hirsch JS, Narasimhan M, Crawford JM, McGinn T, Davidson KW, et al. Presenting Characteristics, Comorbidities, and Outcomes Among 5700 Patients Hospitalized With COVID-19 in the New York City Area. JAMA. 2020.

29. Shi S, Qin M, Shen B, Cai Y, Liu T, Yang F, et al. Association of Cardiac Injury With Mortality in Hospitalized Patients With COVID-19 in Wuhan, China. JAMA Cardiology. 2020.

30. Wang D, Hu B, Hu C, Zhu F, Liu X, Zhang J, et al. Clinical Characteristics of 138 Hospitalized Patients With 2019 Novel Coronavirus–Infected Pneumonia in Wuhan, China. JAMA. 2020;323(11):1061–9.

31. Wu C, Chen X, Cai Y, Xia Ja, Zhou X, Xu S, et al. Risk Factors Associated With Acute Respiratory Distress Syndrome and Death in Patients With Coronavirus Disease 2019 Pneumonia in Wuhan, China. JAMA Intern Med. 2020.

32. Guan WJ, Ni ZY, Hu Y, Liang WH, Ou CQ, He JX, et al. Clinical Characteristics of Coronavirus Disease 2019 in China. The New England journal of medicine. 2020.

33. Yang X, Yu Y, Xu J, Shu H, Xia Ja, Liu H, et al. Clinical course and outcomes of critically ill patients with SARS-CoV-2 pneumonia in Wuhan, China: a single-centered, retrospective, observational study. The Lancet Respiratory Medicine.

34. Organization WH. Clinical management of severe acute respiratory infection when COVID-19 is suspected.

35. Alhazzani W, Moller MH, Arabi YM, Loeb M, Gong MN, Fan E, et al. Surviving Sepsis Campaign: guidelines on the management of critically ill adults with Coronavirus Disease 2019 (COVID-19). Intensive Care Med. 2020.

36. Mo P, Xing Y, Xiao Y, Deng L, Zhao Q, Wang H, et al. Clinical characteristics of refractory COVID-19 pneumonia in Wuhan, China. Clinical infectious diseases: an official publication of the Infectious Diseases Society of America. 2020.

37. Esquinas AM, Egbert Pravinkumar S, Scala R, Gay P, Soroksky A, Girault C, et al. Noninvasive mechanical ventilation in high-risk pulmonary infections: a clinical review. European Respiratory Review. 2014;23(134):427.

38. Hui DS, Hall SD, Chan MT, Chow BK, Tsou JY, Joynt GM, et al. Noninvasive positive-pressure ventilation: An experimental model to assess air and particle dispersion. Chest. 2006;130(3):730–40.

39. Guérin C. Prone ventilation in acute respiratory distress syndrome. European Respiratory Review. 2014;23(132):249.

40. http://clinicaltrials.gov. Awake Prone Positioning to Reduce Invasive VEntilation in COVID-19 Induced Acute Respiratory failurE – Full Text View – http://ClinicalTrials.gov.

41. http://clinicaltrials.gov. Prone Positioning in Awake Patients With COVID-19 Requiring Hospitalization – Full Text View – http://ClinicalTrials.gov.

42. http://clinicaltrials.gov. Prone Position in Patients on High-flow Nasal Oxygen Therapy for COVID-19 (HIGH-PRONE-COVID-19) – Full Text View – http://ClinicalTrials.gov.

